# A Tale of Three (Major) Cities: Trends in Covid-19 Firearm Deaths Pre- vs. Post-Pandemic

**DOI:** 10.1101/2025.04.28.25326367

**Authors:** Cristabella Cardone, Ayana Kilpatrick, Scott Bunker, Riley Ledet, Melody Duran, Carlos Coello-Sanchez, Maria D’Amico, Mark Richman

## Abstract

**Introduction:** Previous studies have identified an increase in the percent of the population dying from firearms during vs. before the COVID-19 pandemic. Such increases have been attributed to a rise in mental health disorders due to medical and financial fear, social isolation, and social unrest. This paper describes in more-depth such changes in the three most-populous counties in the United States: Cook (Chicago), Los Angeles, and New York.

**Methods:** Data was collected from the publicly-available, anonymized/de-identified, and aggregated CDC WONDER database. A chi square test compared rates of firearm deaths by county in 2019 and 2021, with comparisons being made between counties within one year (eg, Cook vs. Los Angeles in 2019) and within a county between years (eg, Cook in 2019 vs. Cook in 2021). We also performed a chi square test to compare changes in the rates of percent of firearms deaths from 2019 to 2021 between the counties.

**Results:** In 2019 and 2021, there were statistically-significant differences in the percent of the population dying from firearms between Cook, Los Angeles, and New York counties (both years: p <0.0001) (Tables 1 and 2). In each county, there was also a statistically-significant increase between 2019 and 2021 in the percentage of population dying from firearms (p <0.0001). (Table 3) However, the rate of increase was not different between the three counties (p = 0.16). That is, the increase in percent of population dying from firearms between 2019 and 2021 in Cook County was not different from that increase in Los Angeles or New York counties.

**Discussion:** Rates of death from firearms increased between 2019 and 2021 in the three largest counties in the United States: Cook, Los Angeles, and New York. The current study validates the findings of prior studies. However, we found the rate of increase was not statistically-significant between the three counties. Numerous causes may underlie the increase, including changes in mental health related to pandemic stressors (physical isolation, unemployment, and actual or fear of poverty^1^) or difficulty accessing mental health services.

## Introduction

The COVID-19 pandemic introduced a plethora of circumstances that increased the appearance of several mental and emotional disorders, including a 3-times increase in anxiety disorders and a 4-times increase in depressive disorder symptoms.^2, 3^ Some stated they experienced this in the first month of the pandemic.^4^ Fear of being fired, and companies actually downsizing, led to financial strain of unemployment, which rose 14% by April 2020 and continued to rise throughout the pandemic. Lack of contact between individuals was a stressor in itself, and provided a barrier to receiving adequate treatment for vulnerable individuals, allowing mental health disorders to mature and reach the point of self-harm and suicidal ideation.^5^ A recent Annals of Emergency Medicine study found firearm violence increased nationally between approximately 25% to 130%, with greater increases in areas with higher social vulnerability.^6^ Between 2020 and 2021, the CDC reported an 8.3% increase in firearm suicides, with this figure increasing throughout the pandemic and never returning to a pre-pandemic value.^7^

Gun violence became a concern during this time as well. During 2020, both the onset of the COVID-19 social distancing order in March 2020 and the social unrest with the George Floyd protests in June 2020, led to concerns and fears about public safety across the U.S.^8^ Firearm purchases in response to fear have increased over the past few decades in the U.S., as the cultural meaning of firearm ownership has shifted from hunting and recreation to personal protection.^9^ At the onset of the social distancing orders, from March to July of 2020, an estimate in excess of 4.3 million firearms were purchased in the US.^10^ Many of these purchasers were first-time gun owners. In 2020, 1.5% of U.S. adults purchased a gun for the first time, an increase of 1.4 million in comparison to the 0.9% of U.S. adults who did so in 2019.^11^

While the COVID-19 pandemic affected everyone in different ways, densely populated urban areas such as Los Angeles, New York City, and Chicago were among the most-affected. One way in which this stress was manifested was an increase in firearm violence. The NYPD reported that, between January 1st and April 4, 2020, there had been 172 shooting incidents, representing an 11.7% increase compared to 2019 and an 18.6% increase compared to 2018. There were 516 shooting incidents reported in Chicago in 2020, a 23% increase compared to 2019 and a 6% increase compared to 2018. In the same study, the LAPD reported a 9.3% decrease compared to 2019 and a 10.3% increase compared to 2018. The psychological, economic, and social impacts of COVID-19 increased the number of recorded violent crimes involving a firearm. An increase in post-COVID firearm-related deaths in Los Angeles, Chicago, and New York City was reported, attributed to many individuals experiencing mental health disorders during the pandemic.^12^

One study, using the CDC FASTER database analyzed firearm-related injuries in ten United States jurisdictions, comparing rates in 2019 vs. those in 2020-2023. This study found an increase from the number of firearm deaths from the years of 2019 to 2023, with the largest increase observed in 2020.^13^

This paper further investigates changes in rates of pre-vs. post-COVID firearm-related death among the three most-populous counties in the United States: Cook (Chicago), Los Angeles, and New York.

## Methods

Data was collected from the publicly-available, anonymized/de-identified, and aggregated CDC WONDER database. We examined the rates of firearm deaths by county from 2019 (pre-COVID-19) to 2021 (during COVID-19). The data was filtered by county: Cook (Chicago), Los Angeles, and New York. There were 7 counties chosen, one in Chicago (Cook County), one in Los Angeles (Los Angeles County), and five in New York City (Bronx County, Kings County, New York County, Queens County, and Richmond County).

When aggregating the total number of deaths for the counties in New York City, Richmond County was eliminated because there were so few firearm deaths that the CDC WONDER database couldn’t give actual data, likely out of concern for researchers or the public to individually identify the victims. Therefore, the number of firearm deaths for Richmond County were not used in this analysis. In addition, the number of firearm deaths were adjusted according to the size of the largest county examined (Los Angeles County), due to the differences in population size.

The population of each county was compared to the number of firearm deaths, and, from that a crude rate per 100,000 was determined. This study examined the changes of rates within each county and compared the differences between the different cities both at each time point (2019, 2021) and between time points (eg, changes in one county from 2019 vs. 2021 vs. changes in another county from 2019 vs. 2021).

A chi square test compared the number of firearm deaths and the population in 2019 and 2021 for the respective counties. An additional chi square test compared the percent of individuals who died from firearms in the respective cities in respect to the overall population in the years 2019 and 2021. For each county, the number of firearm deaths and the total population for the given year was used to determine a crude rate per 100,000. A chi square test was used to compare the same county over two separate years (2019 and 2021). The difference in change of percentage between 2019 and 2021 was calculated. Subsequently, the ratio of changes between years was calculated.

MicrosoftExcel (Redmond, WA; 2021) was utilized for a primary analysis for within-city change and between different cities from 2019 to 2021. Statistical significance was set as a priority at p <0.05.

This project was reviewed and deemed not to meet the definition of research by the Northwell Health Institutional Review Board’s (IRB’s) Human Research Protection Program (HSRD24-0133), indicating that formal IRB approval was not required for this study. All observations were collected in compliance with institutional guidelines for patient privacy and data security.

## Results

Adjusting each county’s population for that of the most-populous county (Los Angeles), in 2019, there were 1,101 firearm deaths in Cook County, 381 in Los Angeles County, and 199 in New York City counties, and in 2021 there were 1,856, 634, and 399 firearm deaths, respectively. (Tables 1 and 2; Figure 1)

**Figure 1.**
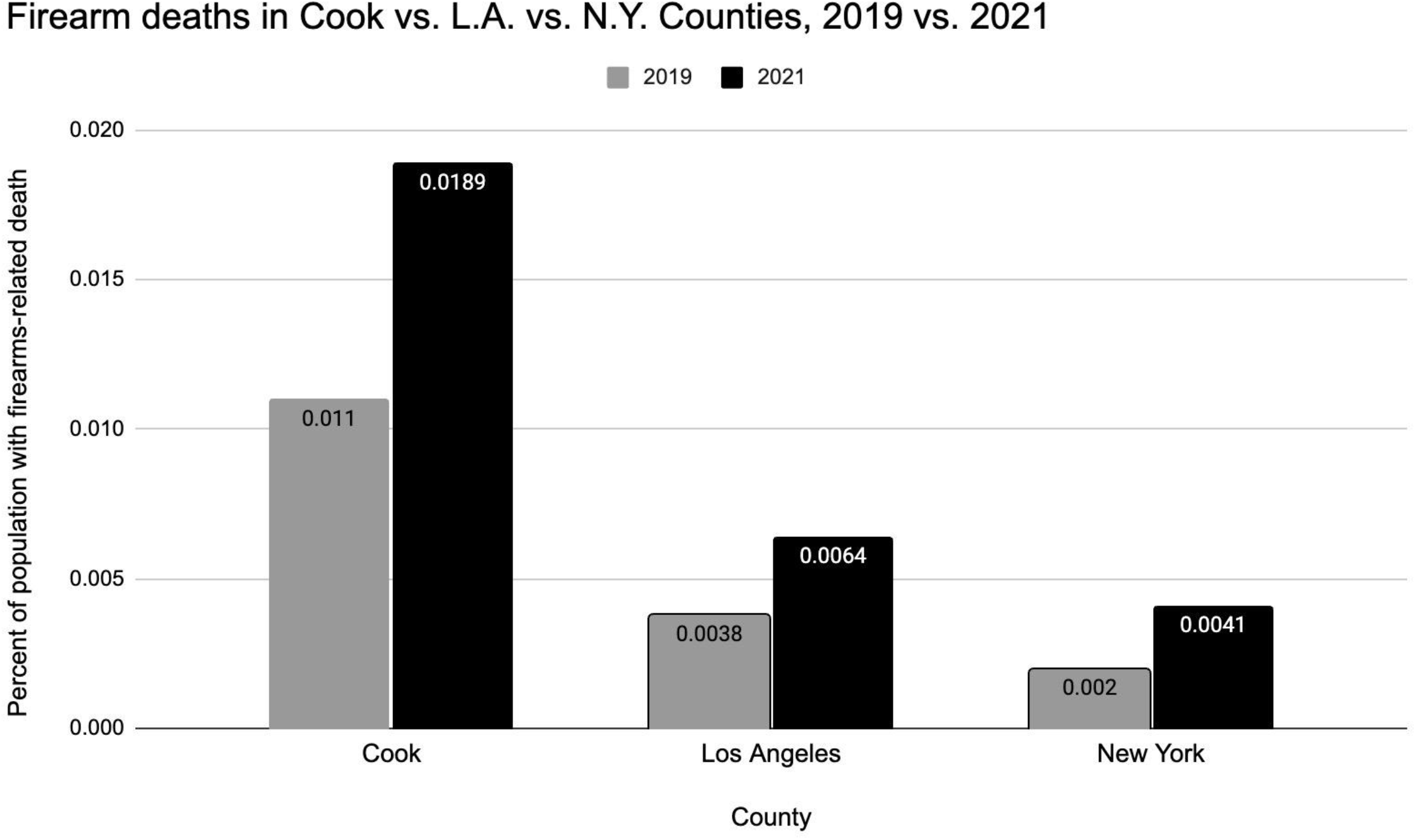
Differences in percent of firearm deaths in 2019 vs. 2021 between Cook County, Los Angeles County, and the aggregated data for New York City counties.

**Table 1.** Firearm deaths in Cook County, Los Angeles County, and the aggregated New York counties in 2019.

| County | Firearm Deaths | Number adjusted for population size of Los Angeles County | Population | Crude Rate (per 100,000) | Percent |
| --- | --- | --- | --- | --- | --- |
| Cook County, IL | 565 | 1,101 | 5,150,233 | 11 | 0.011 |
| Los Angeles County, CA | 381 | 381 | 10,039,107 | 3.8 | 0.0038 |
| New York City counties | 156 | 199 | 7,860,674 | 2 | 0.002 |
| Overall total | 1,102 | 1,681 | 23,526,157 | 4.68 | 0.00468 |
|  |  |  |  | Differences between counties | p <0.00001 |
| Cook County, IL | 977 | 1,856 | 5,173,146 | 18.9 | 0.0189 |
| Los Angeles County, CA | 634 | 634 | 9,829,544 | 6.4 | 0.0064 |
| New York City counties | 324 | 399 | 7,974,019 | 4.1 | 0.0041 |
| Overall total | 1,935 | 2,889 | 23,470,203 | 8 | 0.00829 |
|  |  |  |  | Differences between counties | p <0.00001 |

**Table 2.**
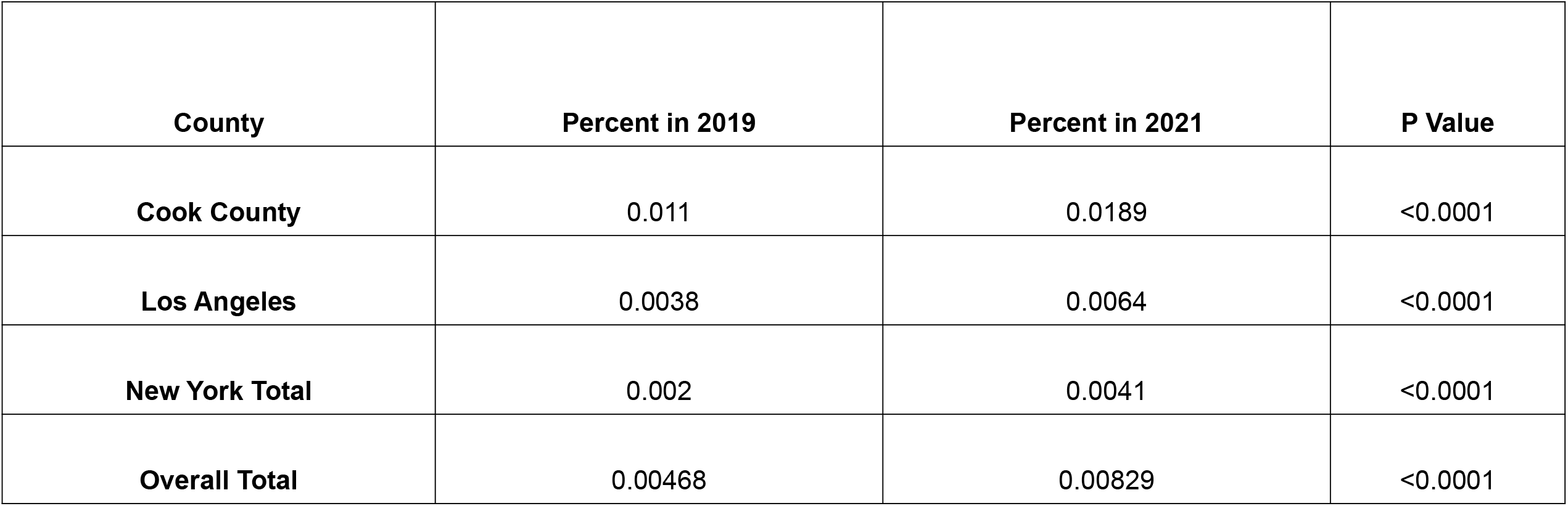
Firearm deaths in Cook County, Los Angeles County, and the aggregated New York counties in 2021.

In 2019 and 2021, there were statistically-significant differences in the percent of the population dying from firearms between Cook, Los Angeles, and the aggregated New York counties (both years: p <0.0001) (Tables 1 and 2). In each county, there was also a statistically-significant increase between 2019 and 2021 in the percentage of population that died from firearms (p <0.0001). (Table 3)

However, the rate of increase was not different between Cook, Los Angeles, and the New York counties (p = 0.16). That is, the increase in percent of population dying from firearms between 2019 and 2021 in Cook County was not different from that increase in Los Angeles or the New York counties.

## Discussion

As previously reported, rates of death from firearms increased in the immediate pre-COVID-19 pandemic time to the time in the early pandemic era. This study provided a deeper, more-granular dive into the data in the United States’ three largest counties: Cook, Los Angeles, and New York. The current study validates prior studies in finding such rates increased. However, the rate of increase was not statistically-significant between the three counties.

Numerous causes may underlie the increase, including changes in mental health related to pandemic stressors (physical isolation, unemployment, and actual or fear of poverty^14^) or difficulty accessing mental health services.

## Data Availability

All data produced are available at https://www.cdc.gov/nndss/infectious-disease/weekly-and-annual-disease-data-tables.html

